# Mindfulness Meditation Training Reshapes Somatomotor Edge-Centric Connectivity Underlying Somatic Awareness: Insights for Healthy and Depressed Individuals

**DOI:** 10.1101/2025.07.29.25331685

**Authors:** Yuwen He, Seng Hang Lei, Weng Si Che, Matthew D. Sacchet, Amy Jing-Wen Yin, Wenyue Gao, Anqi Gu, Natalia Chan, Xinyun Xiao, Jiaze Li, Jieting Chen, Mek Wong, Elvo Kuai Long Sou, Yizhong Shen, Jingju Quan, Weng Ieong Tou, Lin Ian Lio, Chih Chiang Chou, Peng Zhang, Bin Liu, Zhen Yuan

## Abstract

**Background:** Somatic awareness is a fundamental practice for novice mindfulness practitioners. It has been proposed that mindfulness meditation training (MMT) enhances brain function through somatic awareness in both non-clinical and clinical populations. However, the neural signatures underlying this mechanism remain poorly understood.

**Methods:** In our study, we conducted mindfulness interventions in both healthy (51 subjects) and depressed (35 subjects) populations and examined whether somatomotor network (SMN)-related edge-centric functional connectivity (eFC) is modulated by MMT, uncovering the population common and population-specific functioning pathways that may underlie somatic awareness. In addition, we tested whether such SMN-related eFCs can predict the insomnia status across populations.

**Results:** Our findings revealed population-common eFC changes involving the SMN and attentional networks, indicating MMT may modulate the attentional system across populations via somatic awareness. Additionally, we identified population-specific eFC profiles engaging SMN-subcortical networks in the healthy population, suggesting MMT may target more pervasively the automatic processes in the healthy population. The specific eFC profile engaged SMN-default mode network interactions in the depressed population, which may be the functioning mechanism explaining MMT is efficient for depressed individuals. These findings demonstrate the differential neural signatures underlying different focuses of mindfulness via somatic awareness in different populations. Furthermore, support vector regression analysis demonstrated that these altered eFCs significantly predicted improvements in insomnia after MMT in both populations.

**Conclusions:** In summary, our results highlight both shared and distinct neural functioning signatures underlying mindfulness-related mental health improvements via somatic awareness in both the healthy and depressed populations.

## Introduction

The effectiveness of mindfulness in improving both mental and physical health across clinical and non-clinical populations has been well documented [1, 2]. As this evidence base has grown, researchers have increasingly turned their attention to an equally important question: How do mindfulness-based interventions (MBIs) yield such diverse and far-reaching benefits [3, 4]? Neuroimaging studies have proposed that several large-scale functional networks including the default mode network (DMN), salience network (SN), and frontoparietal network (FPN) are the key neural substrates underlying mindfulness [5, 6]. In addition, emerging evidence highlights the involvement of fronto-limbic circuits[7], along with the insula’s role [8] and the cingulate cortex [9]. While these findings underscore the importance of higher-order associative and limbic regions, the potential contributions of somatosensory cortices to mindfulness remain underexplored [10]. More importantly, this gap is particularly striking given that somatic awareness practices, such as body scans and breath-focused attention, are foundational to many mindfulness training programs, particularly for beginners [11, 12]. These practices cultivate present-moment awareness of bodily sensations and serve as entry points for developing the attentional focus, self-awareness, emotion regulation, and other key capacities linked to the therapeutic effects of mindfulness meditation [4]. Therefore, somatosensory processing might play a crucial mediating role in the early stages and effectiveness of mindfulness meditation. However, the specific neural mechanism by which somatosensory regions contribute to mindfulness remains poorly understood. The present study aims to address this gap by examining the role of somatomotor network (SMN), including somatosensory cortices, in MBIs.

SMN consists of the postcentral regions (somatosensory regions), superior temporal regions, premotor and paracentral areas, as well as parts of the insula and superior parietal lobule [13]. These regions maintain extensive connections with both frontal and subcortical brain structures [14] positioning them as potential key mediators in mindfulness-related processes. For example, mindfulness practices have been shown to enhance somatosensory attention [10, 15] and improve perceptual decision-making [16]. Additional magnetoencephalography studies indicate that mindfulness practice enhances local alpha modulation [17], with somatosensory regions playing a central role in these neural changes. Meanwhile, advanced meditation has been related to altered brain network activity and connectivity of somatomotor networks including during advanced concentrative absorption meditation which includes experiences of complete cessation of bodily awareness [18, 19]. Further, [10] emphasized that mindfulness practice often begins with focused attention on bodily sensations, highlighting the importance of somatic awareness. To investigate how mindfulness meditation training (MMT) modulates connectivity between somatosensory regions and other brain networks, SMN-related edge-centric functional connectivity (eFC) were constructed in this study to uncover the neural substrates involved in the functioning mechanisms of mindfulness.

The eFC is a recently developed neuroimaging method that captures intricate neural encoding and processing patterns by examining connectivity between pairs of edges (i.e., connections between brain nodes) [20]. This approach reveals rich information about neural communication and has been shown to reliably involve sensorimotor and attentional networks [20]. Recent work also illustrated eFC’s utility in studying cognitive functions [21], neurological disorders [22], and even its heritability in relation to clinical symptoms [23]. Consequently, eFC is an ideal tool for exploring how MMT modulates SMN-related functional connectivity, offering deeper insights into the neural signatures underlying how MMT functions via somatic awareness.

Meanwhile, MBIs have demonstrated benefits for both clinical and non-clinical populations, though their underlying neural mechanisms might vary depending on the functioning pathways and differential psychopathological sources. In the healthy population, mindfulness training has been associated with reduced stress, improved sleep quality, and enhanced emotion regulation [24–26]. Among clinical groups, such as individuals with major depressive disorder (MDD), mindfulness practices have been shown to alleviate symptoms, including depressive symptoms, duration of relapse, and improve cognitive function [27, 28]. Among them, MBIs could improve sleep quality and attentional control in both the healthy population and MDD. On the other hand, MBIs in general population may bring more emphasis on stress reduction, while MBIs in MDD may focus more on the improvement of depressive symptoms. Such discrepancies of MBIs in different populations may signify their different functioning neural pathways. Uncovering the population-common and population-specific functioning pathways can provide insights into the functioning mechanism of MBIs and offer guidance to develop personalized MBIs for different populations. What’s more, it can also facilitate using neuromodulation approaches to enhance mindfulness practice effects, which is an emerging field in mindfulness research [29].

Insomnia is common among various psychiatric conditions, including MDD, schizophrenia, and so on [30]. It was demonstrated MBIs can improve sleep quality in both the healthy population and MDD. Meanwhile, somatic awareness is a foundation of mindfulness practice, it would be reasonable to infer that somatic awareness may contribute significantly to improving sleep quality. In our study, we would like to examine whether the SMN-related eFCs can predict the sleep status in both the healthy population and MDD. Such a test can help us to examine the importance of SMN-related eFCs in the improvement of symptoms across different populations, which may demonstrate the importance of somatic awareness in the functioning mechanism of MBIs.

In this study, we hypothesized that MMT would modulate SMN-related eFC networks. The resulting changes in SMN-related eFCs can serve as neural signatures underlying how mindfulness achieves its therapeutic effects through somatic awareness in both healthy and MDD individuals. Accordingly, two MMT programs were conducted in healthy and MDD populations separately to examine our hypotheses. Specifically, it is expected from the present study that: 1) Mindfulness training regulates SMN-related eFCs, reflecting on the functioning mechanism of mindfulness meditation via somatic awareness, 2) both population-common and population-specific eFC profiles are modulated by mindfulness meditation, explaining its common and differential functioning effects across populations, and 3) these SMN-related eFCs can predict improvements in mental health outcomes, such as insomnia, in the healthy and MDD populations. By investigating these dynamics, we aim to elucidate how MMT reshapes brain connectivity to improve mental health, offering insights into its neural mechanisms in both healthy and clinical populations.

## Methods

### Participants and Study Protocol

MMT was conducted for the healthy population and the MDD population separately. In the healthy population, forty participants were randomly assigned to a mindfulness (MD) group, while twenty-five participants were assigned to a non-treatment control (NC) group (Figure 1. A). After taking first set of tests (Figure 1. B, T1), Healthy participants in the MD group took a 4-week brief mindfulness workshop for college students to enhance stress management [31], while participants in the NC group received no intervention during this period. At the two-month follow-up, all healthy participants were invited to take the second set of tests (Figure 1. B, T2).

**Figure 1.**
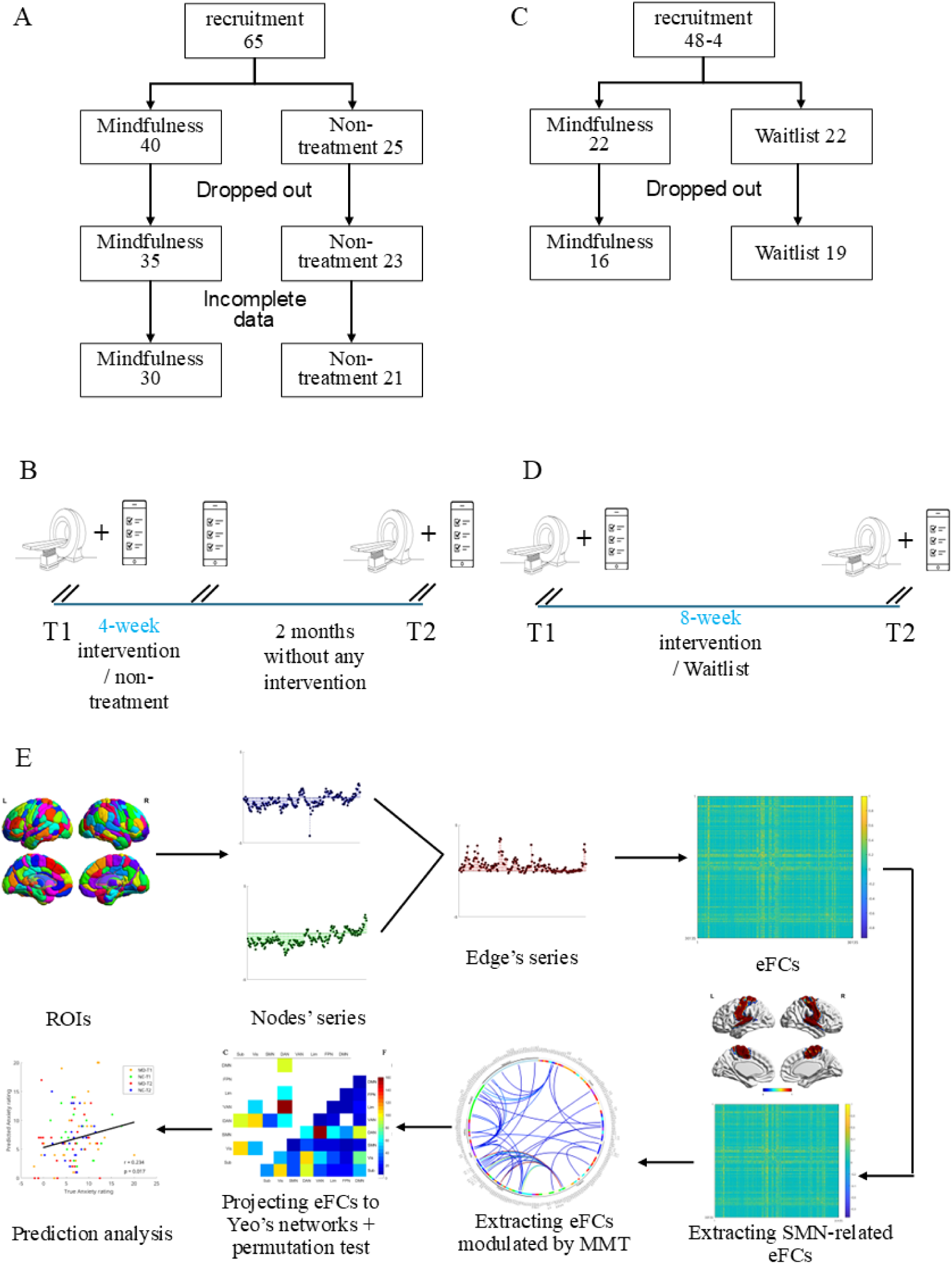
The flow chart of participant recruitment and data analysis. A: The flow chart of participant recruitment in the healthy population; B: The experiment procedures for participants in healthy population: all participants took MRI scans and filled online questionnaires at baseline (T1, pre-intervention); the participants in the MD group were assigned to take a 4-week mindfulness workshop, while the participants in the NC group did not take any intervention during this period; immediately after the 4-week, all participants were invited to take online questionnaires; at the two months follow-up (T2, post-intervention), all participants were invited to take MRI scans and fill the same online questionnaires; C: The flow chart of the recruitment of patients with MDD; D: the experiment procedures for participants in the MDD population: all participants were invited to take MRI scans and online questionnaires at the baseline (pre-intervention, T1); participants in the MD group took part in an 8-week MBCT workshop, while participants in the WL group did not take any particular intervention during this period; 8 weeks later (post-intervention, T2), all participants were invited to take MRI scans and online questionnaires; E: data analysis flow: Brain regions defined by the Brainnetome atlas were used as regions of interest (ROIs) to extract node-wise average time series. Edge-wise time series were then generated by computing the element-wise (dot) product of the time series from each pair of nodes. Pearson correlation coefficients were calculated between all pairs of edge-wise time series to construct the edge functional connectivity (eFC) matrix. Sensorimotor network (SMN)–related eFCs were identified as those containing at least one node within the SMN. Repeated-measures ANOVA was conducted to identify eFCs showing significant group (MD vs. WL) × time (T1 vs. T2) interaction effects. These significant eFCs were subsequently projected onto Yeo’s large-scale functional networks, and permutation tests were performed to determine whether the number of significant eFCs within each network category exceeded chance levels. Finally, the identified significant eFCs were used to predict insomnia severity using support vector regression. Notes: MD: mindfulness; NC: non-treatment; WL: waitlist; ANOVA: analysis of variance.

In the MDD population, twenty-two participants with MDD were randomly assigned to an MD group, taking an 8-week MBCT workshop in addition to their usual treatment, while another twenty-two participants with MDD were assigned to a waitlist (WL) group, receiving no additional intervention apart from their usual treatment during the 8 weeks. All participants with MDD took a set of tests before and after the 8-week period (Figure 1. D).

Finally, 30 healthy participants were left in the MD group, and 21 healthy participants were left in the NC group; 16 participants with MDD were left in the MD group, and 19 participants with MDD were left in the WL group, after removing participants who dropped out early or had incomplete data.

### Clinical Measurement

The Generalized Anxiety Disorder Scale (GAD-7) [32] was used to assess the anxiety level in the healthy population. The Beck depression inventory (BDI) [33] was used to assess the depressive symptoms in the patients with MDD. The insomnia severity index was used to evaluate the insomnia severity in both populations [34].

### Regions of Interest

The Brainnetome atlas was developed based on the structural and functional connectivity profiles of each brain voxel [35]. We took 246 subregions in this atlas for our initial regions of interest (ROIs). In later analysis, the ROIs were assigned to specific Yeo’s resting networks [13], including the subcortical network (Sub), visual network (Vis), SMN, dorsal attention network (DAN), ventral attention network (VAN), limbic network (Lim), frontoparietal network (FPN), and default mode network (DMN).

### Calculation of the Edge-centric Functional Connectivity

Image acquisition and preprocessing procedures were detailed in the supplementary methods. We calculated the edge-centric functional connectivity (eFC) according to the procedures proposed by [20]. The time series in each ROI was extracted and z-scored, and then the dot product of each pair of ROIs was used to index the edge time series of the pair of ROIs. For each pair of edges, the eFC was calculated with Pearson’s correlation with the time series of these two edges.

### Statistical Analysis

In our current study, we focused on the SMN-related eFCs. We extracted these eFCs that at least involved a brain region in the SMN. To reveal the eFCs that were modulated by MMT, we performed repeated-measure ANOVA analysis in each population sample for each eFC. It was followed by counting the eFCs, showing a significant group × time interaction effect (*p* < 0.001 or *p* < 0.01). We then grouped these significant eFCs into categories according to Yeo’s network. For example, if an edge included a region in the SMN and another region in the DMN, then this edge would be classified as category SMN-DMN; then if an eFC included two edges, one edge was classified in SMN-DMN, another was in SMN-DAN, this eFC would be grouped in the eFC category SMN-DMN-SMN-DAN. In addition, we also counted the frequency of a brain region involved in the eFCs showing significant interaction effects in the two data samples.

### Permutation test

We used a random permutation algorithm built in MATLAB to shuffle the significant statistical p-values of all eFCs in the abovementioned repeated-measurement ANOVA. In each permutation, the abovementioned frequency of counted significant eFCs belonging to each category or involved in each ROI was calculated and recorded. These permutation procedures were repeated 10,000 times to generate a null hypothesis probability distribution for each eFC category. We then regarded the eFC category with frequency value higher than 95% of simulated frequences as a reliably significant result, showing reliable modulation effects from MMT

### Support vector regression analysis

In the healthy and MDD populations, we included the eFCs with significant interaction (*p* < 0.001 or *p* < 0.01) to a support vector regression model following procedures used by previous studies [36, 37] to predict clinical measurements significantly improved by mindfulness meditation training. We conducted this multivariate prediction analysis with the Spider toolbox with a linear kernel function (C=1), epsilon was set to 0.1, and 10-fold cross-validation was used to estimate the prediction power. Meanwhile, we also adopted a bootstrapping method to determine the contribution of each eFC to the predictive results. We categorized the eFCs as significant contributors when their corresponding Bootstrapping *p* < 0.005.

## Results

### Demographic characteristics and behavioral results

Healthy individuals in the MD and NC groups were comparable in age, gender, and education (Table 1). Individuals with MDD in the MD and WL groups were comparable in age, gender, education, baseline severity of depressive symptoms, first-episode age, duration of disease, and medication status (Table 1).

**Table 1.** Demographic data in the healthy population.

| Variables | Mindfulness group | Non-treatment<br>\Waitlist group | Chi-square/<br>t-test |
| --- | --- | --- | --- |
| **Healthy population**<br>(Number of participants) | **30** | **21** |  |
| Age (SD) | 23.83(3.56) | 23.86(2.39) | $p = 0.98$ |
| Gender(M/F) | 5/25 | 4/17 | $p = 0.82$ |
| Education (U/M/P) | 10/14/6 | 2/14/5 | $p = 0.14$ |
| Ethnicity (Han/Others) | 28/2 | 19/2 | -- |
| **MDD population**<br>(Number of participants) | **16** | **19** |  |
| Age (SD) | 30.44(6.17) | 29.10(6.56) | $p = 0.54$ |
| Gender(M/F) | 4/12 | 3/16 | $p = 0.49$ |
| Education (U/M/P) | 2/6/8 | 4/9/6 | $P = 0.52$ |
| Baseline Becker<br>depression | 19.47±12.96 | 21.21±9.94 | $P = 0.66$ |
| Ethnicity (Han/Others) | 34/0 | 21/0 | ---- |
| Age of first onset | 26.06±6.96 | 23.94±6.10 | $P = 0.37$ |
| Duration of disease | 3.75±3.70 | 4.06±2.59 | $P = 0.78$ |
| Medicine(Yes/No) | 13/3 | 14/5 | $P = 0.60$ |

Note: (M/F): Male/Female; (U/M/P): undergraduate/Master/Ph.D. SD: standard deviation.

MMT improved anxiety and insomnia in the healthy individuals in the MD group (Figure S1), while these effects were not observed in the NC group. MMT improved insomnia but did not improve depressive symptoms in individuals with MDD in the MD group (Figure S2).

### The eFCs modulated by mindfulness meditation training in the healthy population

As shown in Figure 2. A & D, widespread eFCs were modulated by MMT in the healthy population (*p* < 0.001 or *p* < 0.01). Most of these SMN-related eFCs, showing group × time interaction, engaged brain regions in the inferior frontal gyrus (IFG), cuneus, medial, superior frontal gyrus (SFG), and posterior temporal cortex (Figure 2. B & E, Table S4). Figure 2. C & F show the network-related eFCs modulated by MMT corresponding to two threshold sets (*p* < 0.001 or *p* < 0.01; Tables S5 & S6). After projecting these eFCs into Yeo’s large-scale brain networks and performing a permutation test in both threshold sets, it was evident that MMT mainly modulated eFCs in SMN-subcortical and SMN-attentional networks (DAN and VAN) related categories across different thresholds (highlighted in orange/red squares in Figure 2. C & F), indicating that MMT may significantly influence the bottom-up automatic process and the attentional process via somatic awareness in the healthy population.

**Figure 2.**
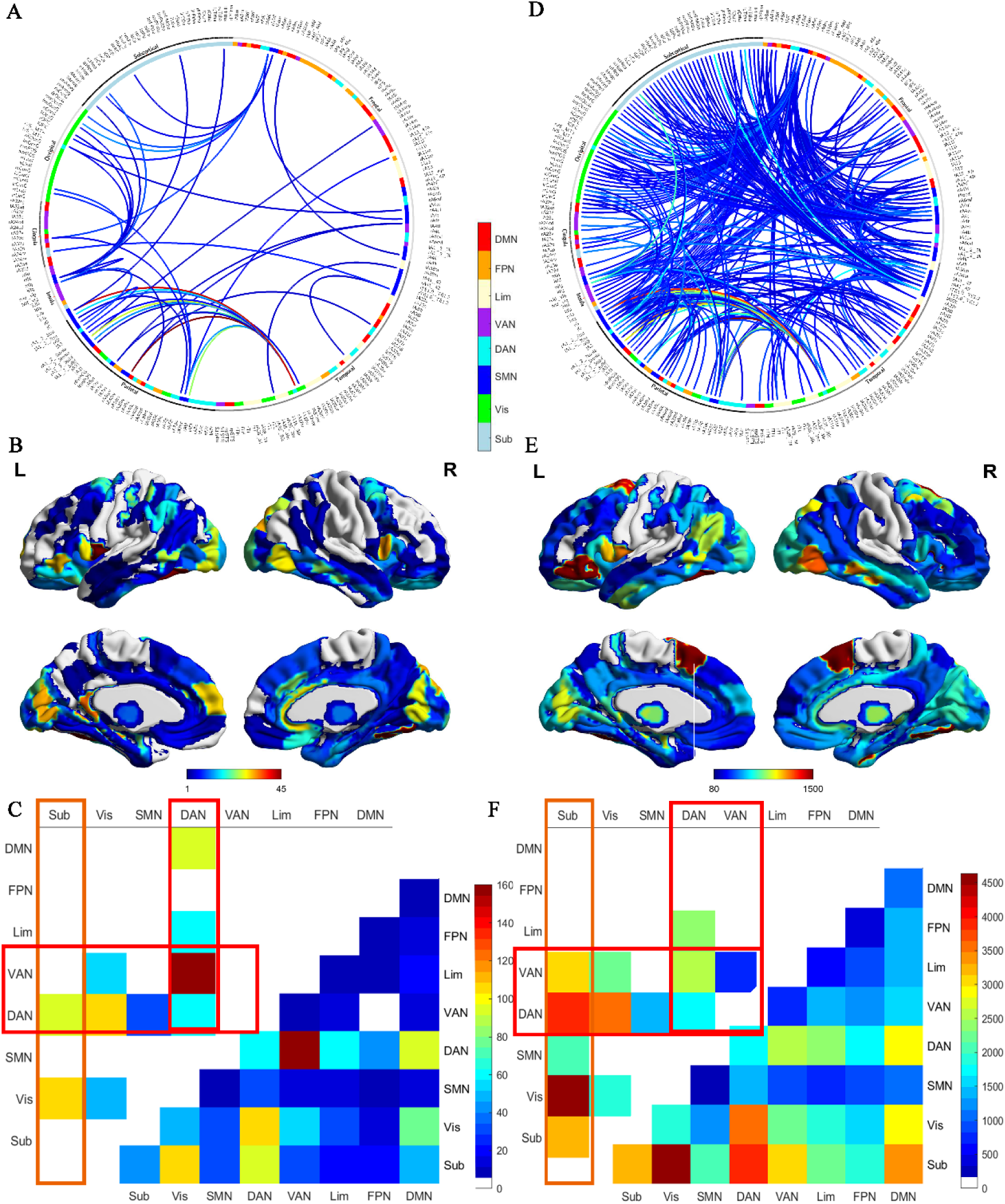
The eFC profile in the healthy population showing a significant group × time interaction. A\D: the frequency of edges involved in eFCs showing a significant group × time interaction with *p* < 0.001 (A) or *p* < 0.01 (D); warmer color of edges indicates higher frequency of engagement; B\E: the frequency of brain regions involved in eFCs showing a significant group × time interaction with *p* < 0.001 (B) or *p* < 0.01 (E); C\F: the frequency of eFCs, showing significant interaction with p < 0.001 (C) or p < 0.01 (F), summarized in network categories (of note, the x- and y-axes are SMN-related edges, i.e., Sub indicates an edge between SMN and subcortical network); the upper triangular parts display frequencies survived a permutation test (the eFC categories highlighted in orange squares are healthy population-specific, while those highlighted in red squares are similar across populations); the lower triangular parts display frequencies without correction. Notes: Sub: subcortical network; Vis: visual network; SMN: somatomotor network; DAN: dorsal attention network; VAN: ventral attention network; Lim: limbic network; FPN: frontoparietal network; DMN: default mode network.

### The eFCs modulated by mindfulness meditation training in the MDD Population

As shown in Figure 3. A & D, diverse eFCs were modulated by MMT in patients with MDD (*p* < 0.001 or *p* < 0.01). Most of these SMN-related eFCs, showing group × time interaction, engaged brain regions in the bilateral middle frontal gyrus (MFG), interior parietal lobule, temporoparietal junction, IFG, cuneus, right posterior cingulate cortex (PCC), and anterior cingulate cortex (ACC), right precuneus, and medial frontal cortex (mPFC) (Figure 3. B & E; Table S4). Figure 3. C & F show the network-related eFCs modulated by MMT corresponding to two threshold sets (*p* < 0.001 or *p* < 0.01; Tables S9 & S10). After the permutation test at the network category level in both threshold sets, it was observed that MMT mainly modulated eFCs in SMN-DMN, and SMN-attentional networks related categories. Similar to healthy population, MMT modulated widespread eFCs engaging SMN and attentional networks, which may be population common neural signatures underlying MMT effects. Of note, widespread eFCs engaging edges in SMN-DMN categories, including eFCs in the SMN-DMN-SMN-Sub, SMN-DMN-SMN-Vis, SMN-DMN-SMN-VAN, SMN-DMN-SMN-FPN, and SMN-DMN-SMN-DMN categories, were reliably modulated by MMT in both threshold sets (*p* < 0.001 or *p* < 0.01), indicating that MMT may influence widespread DMN-related functions via somatic awareness in the MDD population.

**Figure 3.**
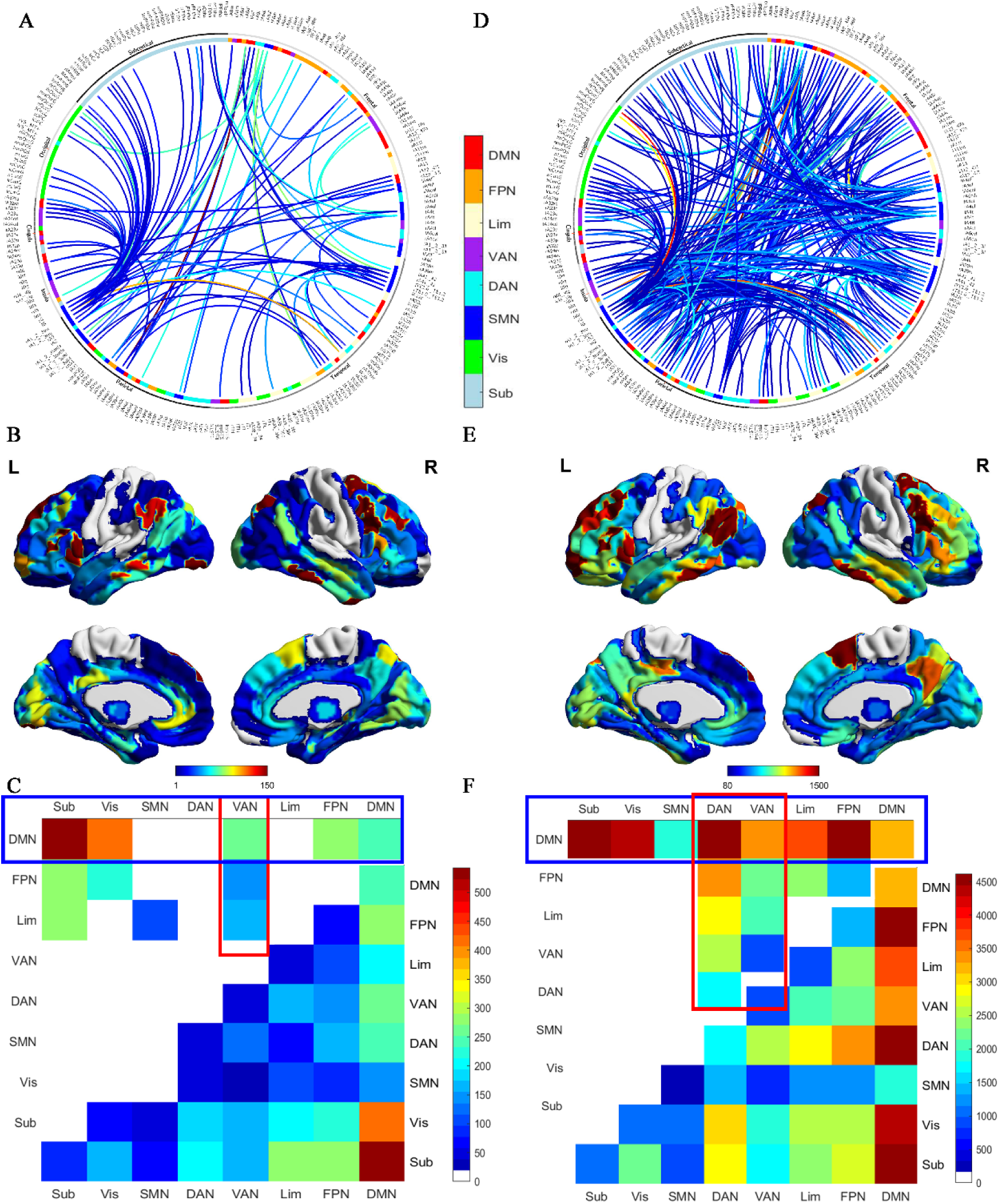
The eFC profile in the MDD population showing a significant group × time interaction. A\D: the frequency of edges involved in eFCs showing a significant group × time interaction with *p* < 0.001 (A) or *p* < 0.01 (D); warmer color of edges indicates higher frequency of engagement; B\E: the frequency of brain regions involved in eFCs showing a significant group × time interaction with *p* < 0.001 (B) or *p* < 0.01 (E); C\F: the frequency of eFCs, showing significant interaction with p < 0.001 (C) or p < 0.01 (F), summarized in network categories (of note, the x- and y-axes are SMN-related edges, i.e., Sub indicates an edge between SMN and subcortical network); the upper triangular parts display frequencies survived a permutation test (the eFC categories highlighted in blue squares are MDD population-specific, while those highlighted in red squares are similar across populations); the lower triangular parts display frequencies without correction. Notes: Sub: subcortical network; Vis: visual network; SMN: somatomotor network; DAN: dorsal attention network; VAN: ventral attention network; Lim: limbic network; FPN: frontoparietal network; DMN: default mode network.

### The prediction results of insomnia with eFCs in the healthy population

As shown in Figure 4. A, the support vector regression prediction model revealed that eFCs (in the set of p < 0.001) can significantly predict the insomnia severity in the healthy population (r=0.28, *p* = 0.004), indicating the eFCs modulated by MMT may be involved in the improvement of insomnia in the healthy population. The other set of eFCs (*p* < 0.01) cannot significantly predict any clinical measurements. The edges of eFCs in the SMN-Vis, SMN-VAN categories showed evident contributions in predicting insomnia (Figure 4. B, Bootstrapping p < 0.005; Table S8).

**Figure 4.**
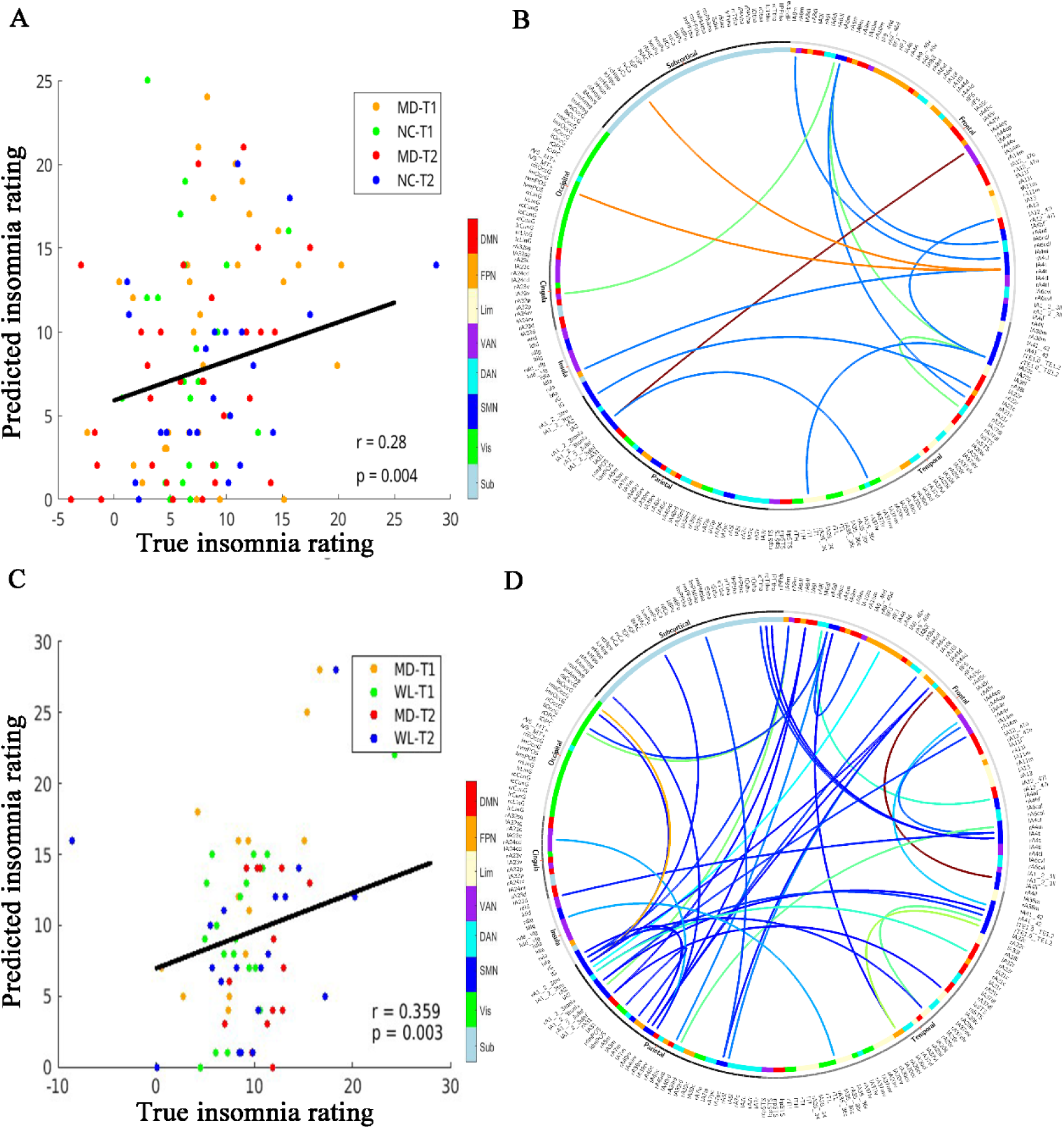
The prediction results of insomnia with eFCs, showing significant group × time p < 0.001, in the healthy and MDD populations. A: the association between true insomnia and predicted insomnia in the healthy population; B: the frequency of edges involved in eFCs having significant contributions to predicting insomnia (p < 0.005) in the healthy population; warmer colors indicate higher frequency of edges; C: the association between true insomnia and predicted insomnia in the MDD population; D: the frequency of edges involved in eFCs having significant contributions to predicting insomnia (p < 0.005) in the MDD population; warmer colors indicate higher frequency of edges. Notes: MD: mindfulness; NC: non-treatment; WL: waitlist; T1: pre-intervention; T2: post-intervention; Sub: subcortical network; Vis: visual network; SMN: somatomotor network; DAN: dorsal attention network; VAN: ventral attention network; Lim: limbic network; FPN: frontoparietal network; DMN: default mode network.

### The prediction results of insomnia with eFCs in the MDD Population

The support vector regression prediction model revealed that eFCs (in the set of *p* < 0.01) can significantly predict the insomnia severity in patients with MDD (Figure 4. C, r = 0.36, *p* = 0.003). The edges of eFCs in widespread categories, including SMN-DAN, SMN-VAN, SMN-Vis, and SMN-DMN, contribute significantly to predicting insomnia (Figure 4. D, Bootstrapping *p* < 0.005; Table S11), indicating the eFCs modulated by MMT may be involved in the improvement of insomnia in patients with MDD.

## Discussion

Our study identified both shared and population-specific profiles of SMN-related eFCs underlying MMT effects in the healthy and MDD populations. These findings provide novel insights into the neural signatures underlying the functional mechanisms of mindfulness—particularly those involving somatic awareness—across different populations. Further analyses revealed that these eFCs significantly predicted insomnia severity in the healthy and MDD populations. These results suggest that the eFCs modulated by MMT may play a critical role in the clinical improvements observed in both populations.

Population-common eFCs modulated by MMT—particularly those involving SMN-DAN and SMN-VAN—suggest a potentially shared neural mechanism across populations. These findings support the hypothesis that enhancement of attentional control via somatic awareness is a core function of diverse mindfulness-based interventions [3, 4, 10]. We found that MMT modulated eFCs engaging SMN and attentional networks in both the healthy and MDD population. A foundational review by Kerr et al. [10] first proposed that somatic awareness in mindfulness practices, such as the Koru Mindfulness curriculum and Mindfulness-Based Cognitive Therapy (MBCT), facilitates the filtering of sensory inputs to the primary sensory cortex and helps organize the flow of sensory information in the brain, thereby enhancing attentional regulation. Extending this hypothesis, our findings reveal specific eFC patterns modulated by MMT that may serve as neural signatures of its mechanisms in attentional functions through somatic awareness across different populations. The dorsal attention network (DAN) plays a central role in attentional control and executive functioning [38], while the ventral attention network (VAN) is involved in salience detection and attentional reorientation [39]. Our observation that mindfulness meditation robustly modulated eFCs in SMN-DAN and SMN-VAN across populations underscores complex interactions among the SMN, DAN, and VAN during mindfulness practice. These interactions may underlie the enhanced attentional control associated with somatic awareness, providing further neural evidence in support of Kerr et al.’s (2013) hypothesis.

The specific eFC profile, regulated pervasively by MMT in the healthy population, involved SMN and subcortical network (Sub). The subcortical network, as defined in our study, includes the amygdala, hippocampus, basal ganglia, and thalamus. These brain regions are fundamental in many automatic processes, including emotion, memory, reward, and perception [40–42]. Uncovering numerous SMN-Sub eFCs modulated by MMT in the healthy population suggests that these connections may underlie the influence of mindfulness on automatic behavioral patterns, basic perception, and salience processing. Thus, these altered eFCs may underlie MMT effects on the ‘autopilot mode’ in healthy individuals [12], encouraging individuals to pay attention to and be aware of what they would perform. Although prior studies have reported changes in functional connectivity involving subcortical regions—such as the amygdala, putamen, and thalamus [43–45]—few have specifically examined the connectivity between the SMN and subcortical networks in the context of mindfulness meditation. Our study provided convincing evidence that MMT would regulate the interactions engaging SMN and attention networks across different populations, which may be the neural signatures underlying MMT effects on attentional control and reducing autopilot behaviors.

Notably, MMT uniquely modulated widespread eFCs in the categories involving edges between SMN and DMN. These changes may underlie improvements in self-referential processes among patients with MDD and provide insight into the functioning mechanism underlying the efficiency of MBCT for patients with MDD. Rumination is characterized in patients with MDD and it is about self-referential processes [46]. Our findings provide neural underpinnings to support the proposal of Kerr et al. (2013) that somatic attention might compete with internally focused rumination and thus efficiently help patients with MDD to improve symptoms. The widespread modulation of SMN-DMN eFCs indicates that self-referential processes are prone to MMT in the MDD population. While many studies have documented DMN alterations associated with MMT [5, 48, 49], relatively few have examined DMN changes following MMT specifically in patients with MDD. Our study provides strong evidence suggesting DMN-related networks are widely modulated by MMT, which may be one of the most important functioning mechanism underlying MMT effects on patients with MDD.

Our further analysis revealed that SMN-related eFCs modulated by MMT in both the healthy and MDD populations significantly predicted changes in insomnia severity. These findings suggest that the modulated SMN-related eFCs may serve as neural signatures underlying the mental health improvements associated with mindfulness meditation across populations. Specifically, in the healthy population, eFCs involving the SMN-VAN, SMN-DMN, SMN-caudate, and SMN-fusiform contributed significantly to predictive performance. This implies that interactions between the SMN and these brain regions may be critical for supporting the mental health benefits of mindfulness in non-clinical individuals. In the MDD sample, eFCs involving the SMN, VAN, SMN-caudate, and SMN-visual cortex (SMN-Vis) were most predictive, highlighting the importance of these interactions in mediating symptom reduction through MBCT in clinical populations. Collectively, these findings indicate that the eFCs identified in our study may represent essential neural mechanisms through which mindfulness meditation, facilitated by somatic awareness, contributes to clinical improvement in both healthy and MDD populations.

Our study has several limitations. First, the mindfulness workshops were tailored for different populations and did not follow identical intervention durations. This difference may have introduced confounding effects in identifying population-specific eFC profiles. However, we posit that intervention duration likely affects the magnitude of connectivity changes more than the pattern of eFC alterations. Second, the sample size for the MDD group was relatively small. This limitation stemmed from a high dropout rate. However, several intervention studies with similar sample size also observed robust interventional effects [57–59], indicating our findings also provide relatively reliable evidence. Future studies with larger clinical samples will be necessary to validate our findings and assess their generalizability.

In conclusion, our study identified population-common changes in eFCs involving SMN and attentional networks after MMT, highlighting the attention regulation in MMT across different populations— Meanwhile, we uncovered population-specific alterations: the healthy population showed pervasive modulations in SMN-Sub-related eFCs, indicating the importance of MMT effects on automatic processes via somatic awareness in the healthy population; whereas the MDD population exhibited widespread changes in SMN-DMN-related eFCs, demonstrating DMN in patients with MDD is prone to MMT. Importantly, these eFCs were associated with improvements in insomnia in both populations. These results offer novel insights into the distinct and shared neural mechanisms underlying mindfulness-based interventions in non-clinical and clinical populations.

## Data Availability

The data can be provided by the University of Macau pending scientific review and a completed material transfer agreement. Requests for the data should be submitted to.

## Acknowledgements

We thank Professor Yuan Zhou’s lab for their valuable suggestions; thank Ms Joanna Kong for her assistance in recruiting participants.

## Notes

### Competing Interest Statement

The authors have declared no competing interest.

### Clinical Trial

ChiCTR2300077225

### Funding Statement

This work was supported by the University of Macau (MYRG2022-00054-FHS, MYRG-GRG2023-00038-FHS-UMDF, and MYRG-GRG2024-00259-FHS), and the Macao Science and Technology Development Fund (FDCT 0014/2024/RIB1).

### Author Declarations

The Ethics committee of the University of Macau gave ethical approval for this work

### Summary of Updates

Revised the manuscript organization and clarify some points missing before

